# Type 2 Diabetes and Risk of Early-Onset Colorectal Cancer

**DOI:** 10.1101/2021.06.02.21257972

**Authors:** Zitong Li, Hanyu Chen, Cassandra D.L Fritz, Xiaobin Zheng, Xiaoyu Zong, Katelin B. Nickel, Andrew Tipping, Long H. Nguyen, Andrew T. Chan, Edward L. Giovannucci, Graham A. Colditz, Margaret A. Olsen, Peter T. Campbell, Nicholas O. Davidson, Ryan C. Fields, Yin Cao

**Affiliations:** Division of Public Health Sciences, Department of Surgery, Washington University School of Medicine, St. Louis, MO, USA; Division of Gastroenterology, Department of Medicine, Washington University School of Medicine, St. Louis, MO, USA; Division of Infectious Diseases, Department of Medicine, Washington University School of Medicine, St Louis, MO, USA; Division of Gastroenterology, Massachusetts General Hospital and Harvard Medical School, Boston, MA, USA; Clinical and Translational Epidemiology Unit, Massachusetts General Hospital and Harvard Medical School, Boston, MA, USA; Broad Institute of MIT and Harvard, Cambridge, Massachusetts; Department of Immunology and Infectious Diseases, Harvard T.H. Chan School of Public Health, Boston, MA, USA; Department of Epidemiology, Harvard T. H. Chan School of Public Health, Boston, MA, USA; Department of Nutrition, Harvard T.H. Chan School of Public Health, Boston, MA, USA; Alvin J. Siteman Cancer Center, Washington University School of Medicine, St. Louis, MO, USA; Department of Population Science, American Cancer Society, Atlanta, GA, USA; Department of Surgery, Washington University School of Medicine, St. Louis, MO, USA

**Keywords:** diabetes, colorectal cancer, early-onset, epidemiology

## Abstract

**Objective:** Early-onset colorectal cancer (CRC) is increasing in many developed countries. Type 2 diabetes mellitus has increased substantially in younger adults; however, its role in early-onset CRC remains unidentified.

**Design:** We conducted a claims-based nested case-control study using IBM® MarketScan® Commercial Database (2006-2015). Incident early-onset CRC diagnosed at ages 18-49 were identified by ICD-9-CM diagnosis code, and the first coded diagnostic pathology date was assigned as the index date. Controls were frequency matched with cases. Type 2 diabetes, stratified by severity, was identified through ICD-9-CM using the Klabunde algorithm. Multivariate logistic regressions were used to estimate odds ratios (ORs) and 95% confidence intervals (Cls).

**Results:** A total of 6001 early-onset CRC and 52104 controls were included. Type 2 diabetes was associated with an increased risk of early-onset CRC (5.0% in cases vs. 3.7% in controls; OR 1.24; 95% CI 1.09 to 1.41). The positive association was more pronounced for uncontrolled (OR 1.37; 95% CI 1.12 to 1.67) or complicated (OR 1.59; 95% CI 1.08-2.35) type 2 diabetes compared to controlled diabetes (OR 1.13; 95% CI 0.94 to 1.36). The positive association was driven by proximal (OR 1.35; 95% CI 1.03 to 1.77) and distal (OR 1.67; 95% CI 1.30 to 2.15) colon cancer but not rectal cancer.

**Conclusions:** Individuals with type 2 diabetes have a higher risk of early-onset CRC, with stronger associations for uncontrolled/complicated diabetes. The rising prevalence of type 2 diabetes among younger adults in the US may partially contribute to the increasing incidence of early-onset CRC.

## INTRODUCTION

Colorectal cancer (CRC) is the second most common cause of cancer-related death in the United States (U.S.).^1^ Due to increased CRC screening and uptake of colonoscopy, the incidence and mortality rates of CRC have declined for several decades among adults aged 50 years and older.^2^ In contrast, the incidence and mortality of early-onset CRC (individuals younger than 50 years) have been increasing since the mid-1990s.^3-5^ During 2012-2016, the incidence of proximal colon, distal colon, and rectal cancer all rose at 1.8% annually among adults younger than 50.^4^ Such an alarming increase in early-onset CRC contributed to a 6 year drop in the median age of CRC diagnosis, from 72 years during 1988-1989 to 66 years during 2015-2016.^5^ Further elucidation of risk factors that contributed to this alarming increase is pivotal.

Thus far, obesity^6^ and sedentary lifestyle^7^ are among the potential contributors to the rise in early-onset CRC, pointing to a possible role of insulin dysregulation. However, the role of type 2 diabetes in early-onset CRC has not been elucidated from large U.S. population-based studies.^8 9^ Although the association between type 2 diabetes and average to late-onset CRC is established,^10-12^ the emerging molecular characteristics of early-onset CRC^13^ support the necessity to revisit such association in a younger population. The link between early-onset CRC with obesity and prolonged sitting, both of which are risk factors for type 2 diabetes,^14 15^ further lend support to such unmet need. Additionally, type 2 diabetes, if etiologically relevant to early-onset CRC, likely contributes to the rising incidence of early-onset CRC due to the paralleled increase of type 2 diabetes^16-23^ and the rise in early-onset CRC in the U.S. and globally.^24^ Specifically in the U.S., between 1988 and 2012, type 2 diabetes has increased from 2.7% to 4.5% for ages 20-44 years, and from 13.3% to 16.2% among ages 45-64.^17^ It is also worth noting that compared with adults with type 2 diabetes at older ages, type 2 diabetes before age 45 appears to be a more aggressive disease with increased risk of requiring insulin.^25^ Therefore, investigating the role of type 2 diabetes in early-onset CRC in a population-based study will likely generate significant insights into the etiology, prevention, and early detection of early-onset CRC.

To address these critical knowledge gaps, we used the IBM® MarketScan® Commercial Database (2006-2015), a longitudinal database that contains individual-level commercial health insurance claims data from over 113 million individuals from all geographic areas of the U.S., to comprehensively examine the association between type 2 diabetes and risk of early-onset CRC.

## METHODS

### Study population

We conducted a nested case-control study of early-onset CRC using the MarketScan Database (2006-2015), a longitudinal, de-identified, individual-level healthcare claims database comprised of more than 113 million commercially insured U.S. adults under age 65.^26^ The database captures information on outpatient and inpatient insurance-reimbursable services, prescription data, type of health plan, and demographic information. Compared with all non-elderly people with employer-sponsored insurance in the U.S., MarketScan enrollees have a similar age and sex distribution.^27^ The Institutional Review Board (IRB) at Washington University in St. Louis decided that this project was not considered to meet federal definitions under the jurisdiction of an IRB and therefore falls outside the purview of the Human Research Protection Office.

### Ascertainment of cases and controls

All adults with an incident diagnosis of CRC between ages 18 and 49 were considered as early-onset CRC and were identified by an International Classification of Diseases, 9^th^ Revision, Clinical Modification (ICD-9-CM) diagnosis code (153.0-153.9, 154.0, 154.1, and 154.8). To reduce false positives, we included only pathology-coded CRC cases and assigned the first diagnostic pathology date as the index date. Confirmed pathology diagnoses are automated into pathology ICD-9-CM diagnosis codes, which were considered accurate in reporting pathology findings.^28^ We restricted our analyses to adults with at least two years of enrollment before the index dates, as well as 90 days of enrollment after the index dates to derive metastatic status. CRCs were further classified into proximal colon (153.0-153.1, 153.4-153.6), distal colon (153.2-153.3, 153.7), unspecified colon (153.8-153.9), and rectal (154.0-154.1) tumor. Metastatic status was imputed using coded diagnosis and/or treatment records for liver/lung metastasis within 90 days of diagnosis.^29^ We excluded CRC patients with any prior/concurrent cancer history (V10.x) or genetic susceptibility to malignant neoplasm (V84.0x), as well as CRC patients with any cancer except non-melanoma skin cancer identified through Cost and Utilization Project’s Clinical Classification Software (HCUP CCS)^30^ within two years before the index dates. The identification of CRC as cases or the other cancers for exclusion for both cases and controls were based on Klabunde *et al*,^31^ which requires at least one inpatient facility claim and/or two outpatient or provider claims 31-365 days apart.

Controls without CRC were frequency matched with cases by up to an 8:1 ratio based on age (18-24 and every five years thereafter), sex (female, male), geographical region (Northeast, North Central, South, West, unknown) due to geographic variations in the incidence of early-onset CRC,^32^ duration of insurance enrollment before index diagnosis (years), and prescription drug coverage (yes, no). Controls were selected to ensure that the distribution of the control index dates matched the distribution of index dates among the cases to account for changes over time. Controls were assigned random index dates and were selected to match the year of the corresponding case’s index date. Controls with genetic susceptibility and personal cancer history were also excluded.

### Ascertainment of type 2 diabetes mellitus

To reduce bias due to increased detection of diabetes and other comorbidities for CRC cases during the workup period but not among controls,^33^ we restricted our exposures and covariates from 91 days to two years before the index dates. Two years before the index dates were chosen to maximize statistical power. More importantly, this time period would allow us to capture almost all patients with diabetes in the target population as 96% of patients with diabetes had at least one diabetes-related appointment with a healthcare provider within a 2-year period.^34^ Type 2 diabetes was defined using ICD-9-CM diagnosis codes (250.00, 250.02, 250.10, 250.12, 250.20, 250.22, 250.30, 250.32, 250.40, 250.42, 250.50, 250.52, 250.60, 250.62, 250.70, 250.72, 250.80, 250.82, 250.90, 250.92) and the Klabunde algorithm.^31^ Type 2 diabetes was further classified as controlled/not stated as uncontrolled without complications (with 250.00, 250.10, 250.20, 250.30 for all the encounters), uncontrolled without complications (with 250.02, 250.12, 250.22, 250.32 in any of the encounters),^35^ or complicated (250.40, 250.42, 250.50, 250.52, 250.60, 250.62, 250.70, 250.72, 250.80, 250.82, 250.90, 250.92).^30^

### Assessment of covariates and other clinical information

We extracted demographic information, including employment status, urban/rural residence, geographical region, health plan, and derived the Charlson Comorbidity Index^36^ without diabetes. We also extracted information on potential confounders, including inflammatory bowel diseases (IBD), obesity, and family history of gastrointestinal cancer between 91 days and 2 years prior to the index dates. Information on fecal occult blood testing (FOBT) and screening/other colonoscopies during the same time period was retrieved. We also obtained information on a list of pre-specified early signs/symptoms for CRC, including gastrointestinal bleeding, abdominal pain, anemia, change of bowel habits, diarrhea, constipation, and weight loss between 91 days and 2 years prior to the index dates.^37^

## Statistical analysis

To evaluate the association between type 2 diabetes and risk of early-onset CRC, multivariable logistic regression models were used to estimate odds ratios (ORs) and 95% confidence intervals (CIs). We first adjusted for matching factors including age (years), sex, duration of insurance enrollment (number of completed years of enrollment), geographical region (Northeast, North Central, South, West, unknown), and prescription drug coverage before the index dates. We then additionally adjusted for full time employment status, residence (urban, rural, unknown), health plan (Preferred Provider Organization, Health Maintenance Organization, others), Charlson Comorbidity Index (continuous), IBD, obesity, family history of gastrointestinal cancer, screening colonoscopy, other colonoscopy, and FOBT. Individuals without geographic region or residence (<2%) were included in the adjustment using missing indicators. To examine the robustness and generalizability of these findings to an asymptomatic population, we conducted sensitivity analyses by restricting to participants without the following: IBD, prior colonoscopy/FOBT, or pre-specified early signs/symptoms of CRC. We also examined whether the association differed according to the severity of type 2 diabetes (controlled, uncontrolled, complicated).

We further examined if the association between type 2 diabetes and early-onset CRC differed according to tumor anatomical site (colon [proximal colon, distal colon, unspecified colon], rectum). We also conducted stratified analyses to evaluate the association among subgroups, including sex, age at the index date (18-45 vs. 46-49 years), birth year (≤1965 vs.>1965), and geographical region (South vs. others). P value for interaction was estimated using a Wald test on the cross-product term of type 2 diabetes and each stratification factor. All the analyses were performed using SAS version 9.4 (SAS Institute, Cary, North Carolina, USA). All the statistical tests were two-sided and P values <0.05 were considered statistically significant.

## RESULTS

A total of 6001 early-onset CRC cases and 52104 controls were included in the analyses (table 1). The mean age of early-onset CRC patients was 43.0 years. Compared with controls, early-onset CRC cases were more likely to have IBD and be coded for obesity. They also had higher rates of colonoscopies other than for screening and FOBT tests before the index dates.

**Table 1.** Characteristics of participants according to case and control status, MarketScan Commercial Database (2006-2015)

|  | **Cases**<br>**(n = 6001)** | **Controls\***<br>**(n = 52104)** |
| --- | --- | --- |
| Age at the index dates (y), mean ± SD | 43.0 ± 5.8 | 42.8 ± 5.8 |
| Female, n (%) | 2922 (48.7) | 25704 (49.3) |
| Geographical region, n (%) |  |  |
| South | 2631 (43.8) | 22624 (43.4) |
| Northeast | 936 (15.6) | 8088 (15.5) |
| North Central | 1380 (23.0) | 11744 (22.5) |
| West | 956 (15.9) | 8824 (16.9) |
| Unknown | 98 (1.6) | 824 (1.6) |
| Duration of insurance enrollment (y), median (IQR) | 3.5 (2.6-5.0) | 3.5 (2.6-4.9) |
| Prescription drug coverage, n (%) | 4951 (82.5) | 43139 (82.8) |
| Full time employed, n (%) | 3075 (51.2) | 27722 (53.2) |
| Residence, n (%) |  |  |
| Urban | 5005 (83.4) | 43937 (84.3) |
| Rural | 902 (15.0) | 7370 (14.1) |
| Unknown | 94 (1.6) | 797 (1.5) |
| Health plan, n (%) |  |  |
| PPO | 3881 (64.7) | 32 672 (62.7) |
| HMO | 776 (12.9) | 7359 (14.1) |
| Other | 1344 (22.4) | 12073 (23.2) |
| Inflammatory bowel disease†, n (%) | 287 (4.8) | 1372 (2.6) |
| Obesity†, n (%) | 428 (7.1) | 2972 (5.7) |
| Family history of gastrointestinal cancer†, n (%) | 75 (1.2) | 510 (1.0) |
| Fecal occult blood test†, n (%) | 452 (7.5) | 3078 (5.9) |
| Screening colonoscopy†, n (%) | 68 (1.1) | 1036 (2.0) |
| Other colonoscopy†, n (%) | 207 (3.4) | 914 (1.8) |
| Charlson Comorbidity Index‡, mean ± SD | 0.1 ± 0.5 | 0.06 ± 0.4 |
| Type 2 diabetes†, n (%) | 302 (5.0) | 1946 (3.7) |
| Anatomical site§, n (%) |  |  |
| Colon | 3846 (64.1) | - |
| Proximal colon | 1156 (19.3) | - |
| Distal colon | 1115 (18.6) | - |
| Unspecified colon | 1575 (26.2) | - |
| Rectum | 2114 (35.2) | - |
| Metastatic status\|\|, n (%) |  |  |
| Non-metastatic | 4560 (76.0) | - |
| Metastatic | 1441 (24.0) | - |

Abbreviations: HMO = Health Maintenance Organization; IQR = interquartile range; PPO = Preferred Provider Organization; SD = standard deviation.
\*Controls were matched based on age (18-24 and every 5 years thereafter), sex (female, male), geographical region (Northeast, North Central, South, West, unknown), duration of insurance enrollment before index diagnosis (years), and prescription drug coverage (yes, no).
†Between 91 days and 2 years before the index date.
‡Charlson Comorbidity Index was calculated without accounting for diabetes.
§A total of 41 cases with more than one anatomical site were excluded.
||Metastatic colon cancer was determined using treatment encounters of liver or lung metastasis within 3 months after the index dates.

Type 2 diabetes was present in 5.0% of early-onset CRC patients, compared with 3.7% among the controls (table 2). In comparison to those without type 2 diabetes, individuals with type 2 diabetes had a 24% increased risk of early-onset CRC (OR, 1.24; 95% CI, 1.09 to 1.41), after adjusting for the matching factors and a range of potential confounders, including full time employment status, residence, type of commercial health insurance, Charlson Comorbidity Index, IBD, obesity, family history of gastrointestinal cancer, personal history of screening colonoscopy, other colonoscopy, and FOBT. The positive association remained similar when we restricted the analysis to individuals without IBD, without family history of gastrointestinal cancer, without previous colonoscopy/FOBT, or without a list of early signs/symptoms of CRC. Secondary analyses of early-onset CRC according to the severity of type 2 diabetes revealed a significant positive association for uncontrolled (OR, 1.37; 95% CI, 1.12 to 1.67) and complicated type 2 diabetes (OR, 1.59; 95% CI, 1.08 to 2.35). However, there was no association between controlled type 2 diabetes and risk of early-onset CRC (OR, 1.13; 95% CI, 0.94 to 1.36).

**Table 2.** Type 2 diabetes and risk of early-onset colorectal cancer

|  | Participants with type 2 diabetes, No. (%) |  | Multivariable adjusted OR (95% CI)* | Multivariable adjusted OR (95% CI)† |
| --- | --- | --- | --- | --- |
|  | Cases | Controls |  |  |
| Any type 2 diabetes |  |  |  |  |
| All participants | 302 (5.0) | 1946 (3.7) | 1.34 (1.19 to 1.52) | 1.24 (1.09 to 1.41) |
| Without IBD | 282 (4.9) | 1862 (3.7) | 1.33 (1.17 to 1.52) | 1.25 (1.09 to 1.42) |
| Without family history of GI cancer | 300 (5.1) | 1928 (3.7) | 1.35 (1.19 to 1.53) | 1.25 (1.10 to 1.42) |
| Without colonoscopy/FOBT | 267 (5.0) | 1731 (3.7) | 1.36 (1.19 to 1.55) | 1.25 (1.09 to 1.43) |
| Without early signs/symptoms‡ | 176 (4.5) | 1278 (3.1) | 1.39 (1.18 to 1.64) | 1.35 (1.14 to 1.59) |
| Type 2 diabetes severity§ |  |  |  |  |
| Controlled | 138 (2.3) | 993 (1.9) | 1.20 (1.00 to 1.44) | 1.13 (0.94 to 1.36) |
| Uncontrolled | 114 (1.9) | 670 (1.3) | 1.47 (1.20 to 1.80) | 1.37 (1.12 to 1.67) |
| Complicated | 32 (0.5) | 146 (0.3) | 1.89 (1.29 to 2.77) | 1.59 (1.08 to 2.35) |

Abbreviations: CI = confidence interval; FOBT = fecal occult blood test; GI = gastrointestinal; IBD = inflammatory bowel disease; OR = odds ratio.
\*Adjusted for matching factors including age (year), sex (female, male), duration of insurance enrollment (year), region (Northeast, North Central, South, West, unknown), and prescription drug coverage (yes/no).
†Additionally adjusted for employment status (full time/others), residence (urban, rural, unknown), health plan (PPO, HMO, others), Charlson Comorbidity Index without diabetes (continuous), and any of the following conditions between 91 days to 2 years before the index dates: IBD (yes/no), obesity (yes/no), family history of gastrointestinal cancer (yes/no), screening colonoscopy (yes/no), other colonoscopy (yes/no), and fecal occult blood test (yes/no).
‡Early signs/symptoms included any of the following conditions between 91 days to 2 years before the index dates: gastrointestinal bleeding, abdominal pain, anemia, change of bowel habits, diarrhea, constipation, and weight loss.
§A total of 18 type 2 diabetes (out of 302) patients among cases and 137 among controls (out of 1946) could not be classified as controlled, uncontrolled, or complicated.

We further evaluated the association between type 2 diabetes and early-onset CRC according to anatomical location of the CRC (table 3). The positive association was largely driven by proximal colon (OR, 1.35; 95% CI, 1.03 to 1.77) and distal colon cancer (OR, 1.67; 95% CI, 1.30 to 2.15) rather than by rectal cancer (OR, 1.13; 95% CI, 0.92 to 1.40).

**Table 3.** Type 2 diabetes and risk of early-onset colorectal cancer according to anatomical site

|  | Participants with type 2 diabetes, No. (%) |  | Multivariable adjusted OR (95% CI)* | Multivariable adjusted OR (95% CI)† |
| --- | --- | --- | --- | --- |
|  | Cases | Controls |  |  |
| **Colon cancer‡** | 204 (5.3) | 1946 (3.7) | 1.43 (1.23 to 1.66) | 1.32 (1.13 to 1.54) |
| Proximal colon | 60 (5.2) | 1946 (3.7) | 1.48 (1.14 to 1.93) | 1.35 (1.03 to 1.77) |
| Distal colon | 72 (6.5) | 1946 (3.7) | 1.72 (1.35 to 2.19) | 1.67 (1.30 to 2.15) |
| Unspecified colon | 72 (4.6) | 1946 (3.7) | 1.19 (0.94 to 1.52) | 1.06 (0.83 to 1.36) |
| **Rectal cancer‡** | 94 (4.6) | 1946 (3.7) | 1.22 (0.99 to 1.50) | 1.13 (0.92 to 1.40) |

Abbreviations: CI = confidence interval; OR = odds ratio.
\*Adjusted for the same set of covariates as model \* in Table 2.
†Adjusted for the same set of covariates as model † in Table 2.
‡A total of 41 cases with more than one anatomical site were excluded.

In stratified analyses, the associations between type 2 diabetes and early-onset CRC according to sex (female vs. male), age (18-45 vs. 46-49 years), and geographic region (South vs. others) were similar and no significant interactions were identified (all P >0.05 for interaction). The association appeared stronger for persons born after 1965 (OR, 1.31; 95% CI, 1.09 to 1.42) compared to those before 1965 (OR, 1.19; 95% CI, 0.99 to 1.42), although no interaction was observed (P = 0.44 for interaction) (table 4).

**Table 4.** Stratified analyses of type 2 diabetes and risk of early-onset colorectal cancer

|  | Participants with type 2 diabetes, No. (%) |  | Multivariable adjusted OR (95% CI) * | P for interaction† |
| --- | --- | --- | --- | --- |
|  | Cases | Controls |  |  |
| **Sex** |  |  |  |  |
| Female | 135 (4.6) | 873 (3.4) | 1.26 (1.04 to 1.52) | 0.92 |
| Male | 167 (5.4) | 1073 (4.1) | 1.23 (1.03 to 1.46) |  |
| **Age at the index date** |  |  |  |  |
| ≤ 45 | 96 (3.3) | 590 (2.3) | 1.28 (1.02 to 1.60) | 0.66 |
| 46-50 | 206 (6.7) | 1356 (5.0) | 1.25 (1.07 to 1.45) |  |
| **Birth year** |  |  |  |  |
| ≤ 1965 | 152 (6.3) | 1005 (5.0) | 1.19 (0.99 to 1.42) | 0.44 |
| > 1965 | 150 (4.2) | 941 (2.9) | 1.31 (1.09 to 1.57) |  |
| **Geographical region** |  |  |  |  |
| South | 155 (5.9) | 994 (4.4) | 1.26 (1.06 to 1.51) | 0.99 |
| Others | 147 (4.4) | 952 (3.2) | 1.23 (1.03 to 1.48) |  |

Abbreviations: CI = confidence interval; OR = odds ratio.
\*Adjusted for the same covariates as the model † in Table 2 without the stratification factor.
†P for interaction was calculated by Wald test using the cross-product term of type 2 diabetes and each stratification factor.

## DISCUSSION

In this nested case-control study leveraging real-world claims data with 6001 early-onset CRC cases, we found that type 2 diabetes was associated with a 24% increased risk of developing early-onset CRC compared to individuals without type 2 diabetes. The positive association, largely observed for proximal and distal colon cancer, remained similar when restricted to individuals without IBD, family history of gastrointestinal cancer, previous colonoscopy/FOBT, or early signs/symptoms of CRC. We also found that this association was more pronounced for uncontrolled or complicated type 2 diabetes compared to controlled type 2 diabetes. Our findings suggest that type 2 diabetes contributes, in part, to the rising incidence of early-onset CRC.

A recent systematic review^38^ estimated that type 2 diabetes was associated with a 27% increased risk of CRC among older individuals. Our study supports a similar association between type 2 diabetes and risk of early-onset CRC. Findings from recent studies attempting to examine the link between type 2 diabetes and early-onset CRC were mixed. While Low *et al* did not observe an association between type 2 diabetes and early-onset CRC,^9^ Khan *et al* reported that in a Swedish nationwide cohort, type 2 diabetes was associated with increased risk of early-onset CRC.^8^ However, only 31 patients with type 2 diabetes were documented prior to early-onset CRC. Moreover, the prevalence of type 2 diabetes (1.4%) in this analytic cohort was significantly lower than that in the US population. With a total of 302 patients with type 2 diabetes preceding 6,001 early-onset CRC cases, our study is among the first with adequate power to provide a reliable effect estimate for the association between type 2 diabetes and early-onset CRC. Notably, our results are likely generalizable to the U.S. population as the prevalence of type 2 diabetes among our controls corresponds to the U.S. national data.^17^ The similar associations observed across strata of sex, birth year, and geographical regions (South, historically described as the “diabetes belt,” ^39^ vs. others) further ensure the generalizability of our findings.

The underlying mechanisms of the association between type 2 diabetes and CRC are not fully understood. Insulin resistance, hyperglycemia, and hyperinsulinemia may play important roles.^40 41^ Our observed positive association for uncontrolled but not for controlled type 2 diabetes, further supports that hyperglycemia/hyperinsulinemia are likely critical to colorectal carcinogenesis. Impaired insulin receptor activation and subsequent defective PI3K signaling pathway could lead to insulin resistance and hyperinsulinemia,^42^ as well as high levels of insulin-like growth factor (IGF) 1.^43^ The insulin-PI3K pathway has been shown to have profound effects on cancer initiation^42^ by stimulating colonic mucosal cell growth and sustaining tumor growth. IGFs are key regulators in signal transduction networks that have important roles in neoplasia.^44^ Uncontrolled hyperglycemia in type 2 diabetes might contribute to DNA damage and aberrant RNA expression that could promote carcinogenesis and cancer progression.^45^ Moreover, a dysregulated immune system in type 2 diabetes patients and chronic inflammation accompanied with elevated cytokine level such as interleukin-6, tumor necrosis factor-α, and C-reactive protein could promote CRC tumorigenesis.^46^ Finally, altered host-microbiota crosstalk^47^ and increased colonic transit time in type 2 diabetes have been linked to dysregulation of bile acid metabolism,^48^ which could also contribute to colorectal carcinogenesis.

It is worth noting that the observed associations between type 2 diabetes and early-onset CRC were evident only for colon but not rectal cancer. Emerging evidence indicates that the molecular features of CRC vary by anatomic subsites. Microsatellite instability (MSI), CpG island methylator phenotype (CIMP), and *BRAF* mutation gradually increase from the rectum to the ascending colon.^49^

Rectal cancers also exhibit more *TP53* mutations and fewer *PIK3CA* mutations or *CTNNB1* mutations.^50^ Given the heterogeneous nature of CRC, it is expected that risk factors also differ by anatomic location. In line with our findings in younger adults, a case-control analysis involving 21744 CRCs from Veterans (median age 68 years) reported that diabetes was associated with 29%, 15%, and 12% increased risk of proximal, distal, and rectal cancer, respectively.^51^ It is hypothesized that IGF1 activates the PI3K/AKT pathway through *PIK3CA* mutation.^42^ The lower frequency of *PIK3CA* mutations in rectal cancer may in part explain the lack of association between type 2 diabetes and rectal cancer. Further elucidations of mechanisms underlying these differential associations are critical for the development of precision prevention strategies.

Our study has several strengths. This large, nested case-control study leveraged longitudinal claims data from close to half of the U.S. adult population. Such an unprecedented sample size provided a unique opportunity to examine the association of interest. This type of examination between type 2 diabetes and early-onset CRC is not otherwise feasible in existing prospective cohort studies or other real-world EHR-based databases, due to the relatively low prevalence of type 2 diabetes and low incidence of early-onset CRC in younger adults. Our rigorous study design restricted CRC cases to those only with confirmed pathology claims, and type 2 diabetes patients were identified through ICD-9-CM coding followed by an established algorithm to maximize reliability. To minimize potential detection bias from patients presenting with signs/symptoms that directly led to a diagnosis of CRC, we leveraged claims data from 91 days to two years before CRC diagnosis and adjusted for a list of variables associated with detection. We also conducted sensitivity analyses to minimize the influence of comorbidities/symptoms that may have led to differential detection of type 2 diabetes among cases and controls.

The study also has a few limitations. First, we were not able to reliably identify the first date of type 2 diabetes diagnosis and thus could not assess the impact of duration of diabetes. However, among the prospective studies that allowed such assessment, there were a limited number of CRC cases with diabetes,^52^ and such investigation is largely infeasible for early-onset CRC. Second, residual confounding could not be ruled out due to limited information on confounders such as smoking, alcohol intake, diet, and physical activity. Studies on the associations between these factors and early-onset CRC are limited. Furthermore, these lifestyle factors are likely modest risk factors for type 2 diabetes and early-onset CRC. Indeed, in analyses among an older population, these factors minimally confound the association between type 2 diabetes and CRC of later-onset.^53^ Third, the MarketScan database does not provide information on race/ethnicity and is restricted to individuals with commercial insurance. Thus, further validation in diverse groups is warranted.

## Conclusions

In this large U.S. claims-based nested case-control study, type 2 diabetes was associated with increased risk of early-onset CRC, suggesting that the rising incidence of early-onset CRC may be partially attributed to the surging prevalence of type 2 diabetes. The more pronounced association for uncontrolled or complicated diabetes further highlights the importance of early detection and intervention of type 2 diabetes at younger ages. Our findings lend support to the promise of type 2 diabetes control as an emerging CRC prevention strategy among younger adults.

## Data Availability

Data were obtained from a third party and are not publicly available.

## Abbreviations

CI: confidence interval
CRC: colorectal cancer
FOBT: fecal occult blood test
GI: gastrointestinal
HCUP CCS: Healthcare Cost and Utilization Project’s Clinical Classification Software
HMO: Health Maintenance Organization
IBD: inflammatory bowel diseases
IQR: interquartile range
OR: odds ratio
PPO: Preferred Provider Organization
SD: standard deviation

## Author contributions

Drs. Li, Chen, Fritz, and Cao had full access to all of the data in the study and take responsibility for the integrity of the data and the accuracy of the data analysis.

Study concept and design: ZL, HC, CF, XBZ, MAO, YC.

Acquisition of data: KBN, AT, MAO, YC.

Analysis and interpretation of data: All authors.

Drafting of the manuscript: ZL, HC, CF, XBZ, YC.

Critical revision of the manuscript for important intellectual content: All authors.

Statistical analysis: ZL, HC, XYZ, YC.

Obtained funding: YC

Administrative, technical, or material support: YC.

Study supervision: YC.

All authors approved the final submitted draft of the manuscript.

